# Public Perceptions of Artificial Intelligence and Robotics in Medicine

**DOI:** 10.1101/2019.12.15.19014985

**Authors:** Bethany Stai, Nick Heller, Sean McSweeney, Jack Rickman, Paul Blake, Ranveer Vasdev, Zach Edgerton, Resha Tejpaul, Matt Peterson, Arveen Kalapara, Subodh Regmi, Nikolaos Papanikolopoulos, Christopher Weight

## Abstract

**Objective:** To understand better the public perception and comprehension with medical technology such as artificial intelligence and robotic surgery. Additionally, to identify sensitivity to, and comfort with, the use of AI and robotics in medicine a in order to ensure acceptability and quality of counseling and to guide future development.

**Subjects and Methods:** A survey was conducted on a convenience sample of visitors to the Minnesota State Fair (n = 264). The survey investigated participant beliefs on the capabilities of AI and robotics in medicine and their comfort with such technology. Participants were randomized to receive one of two similar surveys. In the first a diagnosis was made by a physician and in the second by an AI application in order to compare confidence in human and computer-based diagnosis.

**Results:** The median age of participants was 45 (IQR 28-59), 58% were female (n=154) vs. 42% male (n=110), 69% had completed at least a bachelor’s degree, 88% were Caucasian (n=233) vs. 12% ethnic minorities (n=31) and were from 12 states in the US with most from the Upper Midwest. Participants had nearly equal trust in AI vs. physician diagnoses, however, they were significantly more likely to trust an AI diagnosis of cancer over a doctor’s diagnosis when responding to the version of the survey that suggested an AI could make medical diagnosis (p = 9.32e-06). Though 55% of respondents (n=145) reported they were uncomfortable with automated robotic surgery the majority of the individuals surveyed (88%) mistakenly believed that partially autonomous surgery was already being performed. Almost all (94%) stated they would be willing to pay for an AI to review their medical imaging, if available.

**Conclusion:** Most participants express confidence in AI providing medical diagnoses, sometimes even over human physicians. Participants generally expressed concern with surgical AI, but mistakenly believe it is already happening. As AI applications make their way into medical practice, health care providers should be cognizant of patient misconceptions and the sensitivity that patients have to how such technology is represented.

## Introduction

Recent advances in computer vision and machine learning have introduced a new wave of technologies to enhance the care of patients. Applications such as automatic measurement of spatial features from medical imaging [1], precision medicine [2] and predictive methods describing the potential course of a disease [3] are being actively developed. With such a wide variety of new applications there has been much activity in development of data sets and methods however relatively little research has been done on the public’s perception of artificial intelligence in the care they receive. As a comparison, research into autonomous vehicles has been similarly rapid and much has been written about the public perception of autonomous vehicles and the potential implementation [4][5]. Small scale initial research has been done on the intersection of medicine and AI from a consumer perspective [6]. This research found a negative association with AI and medical care in the minds of consumers. This seems to indicate a tension between the widespread research interest and the public interest in AI.

The technological advancement of AI has paralleled the development and introduction of robotic surgery. Laparoscopic surgery in urology has been on the forefront of new surgical technology with the use of robotics in procedures such as nephrectomy, prostatectomy and cystectomy [7]. Previous studies which examine patients’ comfort and perception of such surgery[8] [9] provide useful precedent when examining patients’ view of AI. A common thread in this research has shown that there is misunderstanding about capabilities and extend of current use, but there is a general optimism and prestige associated with new technologies. Boys et al. found that almost 20% of the respondents believed the robot itself had some degree of autonomy, offering a natural topic in which to investigate individuals’ perceptions of AI in medicine. The aim of this work is to explore public perceptions of AI in medicine, and evaluate relationships between demographic characteristics and disposition toward AI and robotics in the treatment and diagnosis of cancer.

## Subjects and Methods

A questionnaire was developed to investigate attitudes and acceptance of new technologies in medicine such as AI and robotic surgery. The survey was administered to a convenience sample of attendees at the Minnesota State Fair on two days in 2019. Inclusion criteria were individuals 18 years or older who volunteered for and completed our questionnaire. Those who participated were given a nominal prize valued at less than $5. The survey was self-administered digitally on tablets.

Demographic information was collected including information on gender, age, level of highest education, race, ethnicity, occupation and zip code. Additional demographic information relevant to the zip code as recorded in the 2017 American Community Survey [10] was incorporated. These variables include the median household income, a designation of whether the zip code falls in a majority rural county, and the proportion of households that report having a broadband internet connection.

Participants were asked a series of questions relating to the use of robotic surgery and artificial intelligence (AI) applications in the treatment of cancer. The questions fall into four topics:

1. Active Surveillance
2. Autonomous Surgery
3. Conflicting Diagnosis Between a Doctor and AI
4. Consumer Facing Image Reading by AI

Each participant randomly received one of two versions of the survey. The only difference between the two versions was the first question, which (paraphrasing) was either (1) “a radiologist believes there’s a 25% chance your renal mass is cancer” or (2) “an AI believes there’s a 25% chance your renal mass is cancer”. In both cases, the participant was told that the recommended course is Active Surveillance. They were then asked whether they would pursue a second opinion. If a participant responds with “yes” they were given a follow-up question asking at what estimated confidence in benignity would they feel comfortable with active surveillance. Options included: ‘76-80%’, ‘81-85%’, ‘86-90%’, ‘91-95%’, ‘96-99%’ and ‘>99%’.

A second set of questions investigated the participants’ perceptions of robotic surgery. First, they were asked to estimate what proportion of a standard operation involving the surgical robot is performed autonomously, and then they were asked to rate their comfort with robotic surgery on a Likert Scale.

The next question asked the participant to select the diagnosis in which they have the highest confidence between an AI and a doctor when the doctor believes a mass is cancerous while an AI predicts that it is benign. The last question asked if the participant would be comfortable with using a web service that evaluated their medical imaging in real time using an AI algorithm in order to get *a second opinion* whether a mass was cancerous or not. If they indicated that they would be comfortable with such a service, they were then asked what they would be comfortable paying for such a service. Options were ‘$0’, ‘$1-10’, ‘$11-20’, ‘$21-50’, ‘$51-100’ and ‘>$100’.

Responses were captured and stored in a REDCap (REDCap; Vanderbilt University, Nashville, TN, USA) database through the University of Minnesota and then exported for analysis in R version 3.4.4 (R Foundation for Statistical Computing, Vienna, Austria). Multivariate regressions and multivariate logistic regressions were used as appropriate to identify associations between demographic factors and responses. P-values less than 0.05 were considered to be significant. This survey was administered with the approval and oversight of the Institutional Review Board of the University of Minnesota and informed consent was obtained from all participants.

## Results

Responses were collected from 298 participants over two days of data collection. After removing incomplete responses, there were a total of 264 participants included in our study. Demographics of the participants are outlined in Table 1 for both survey versions 1 and 2. Survey response counts by question are found in Table 2.

**Table 1:**
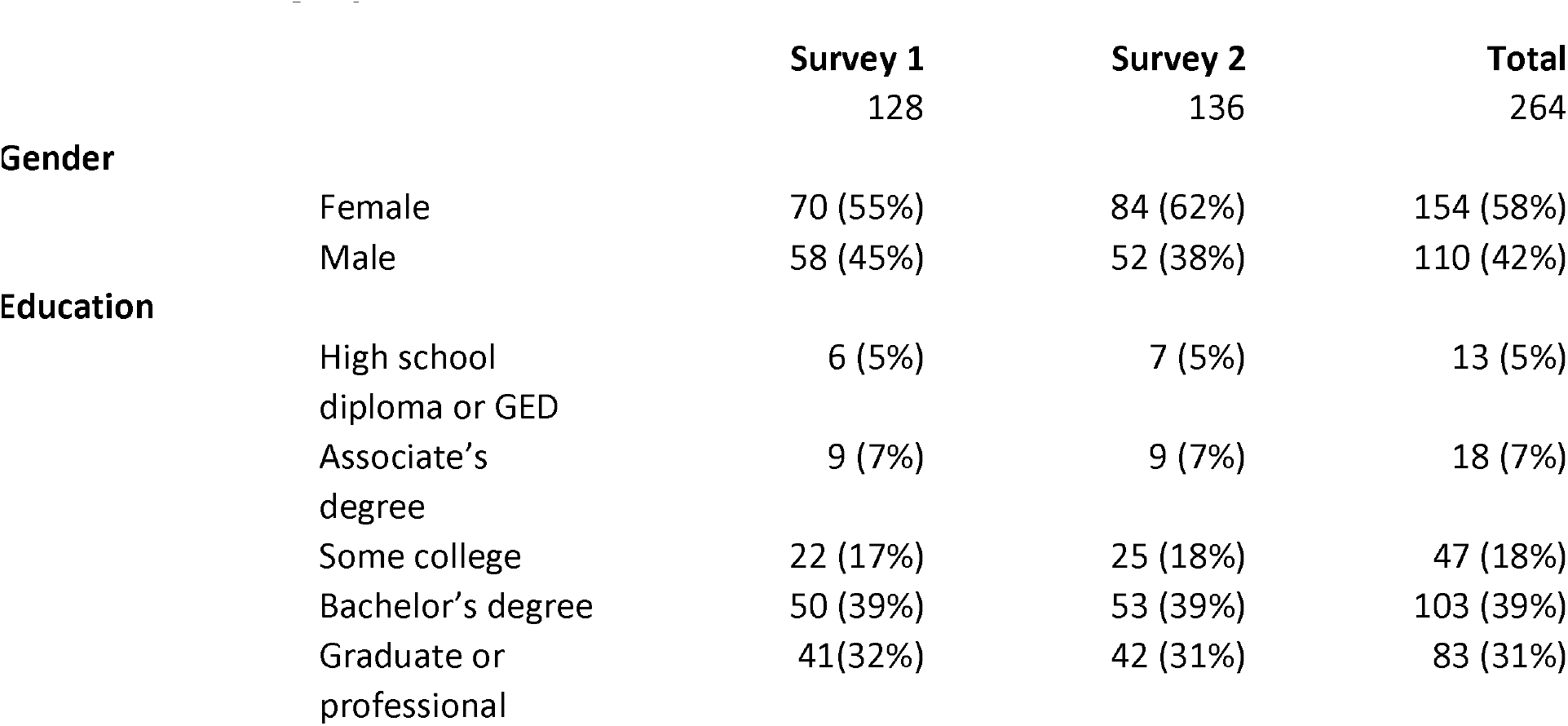
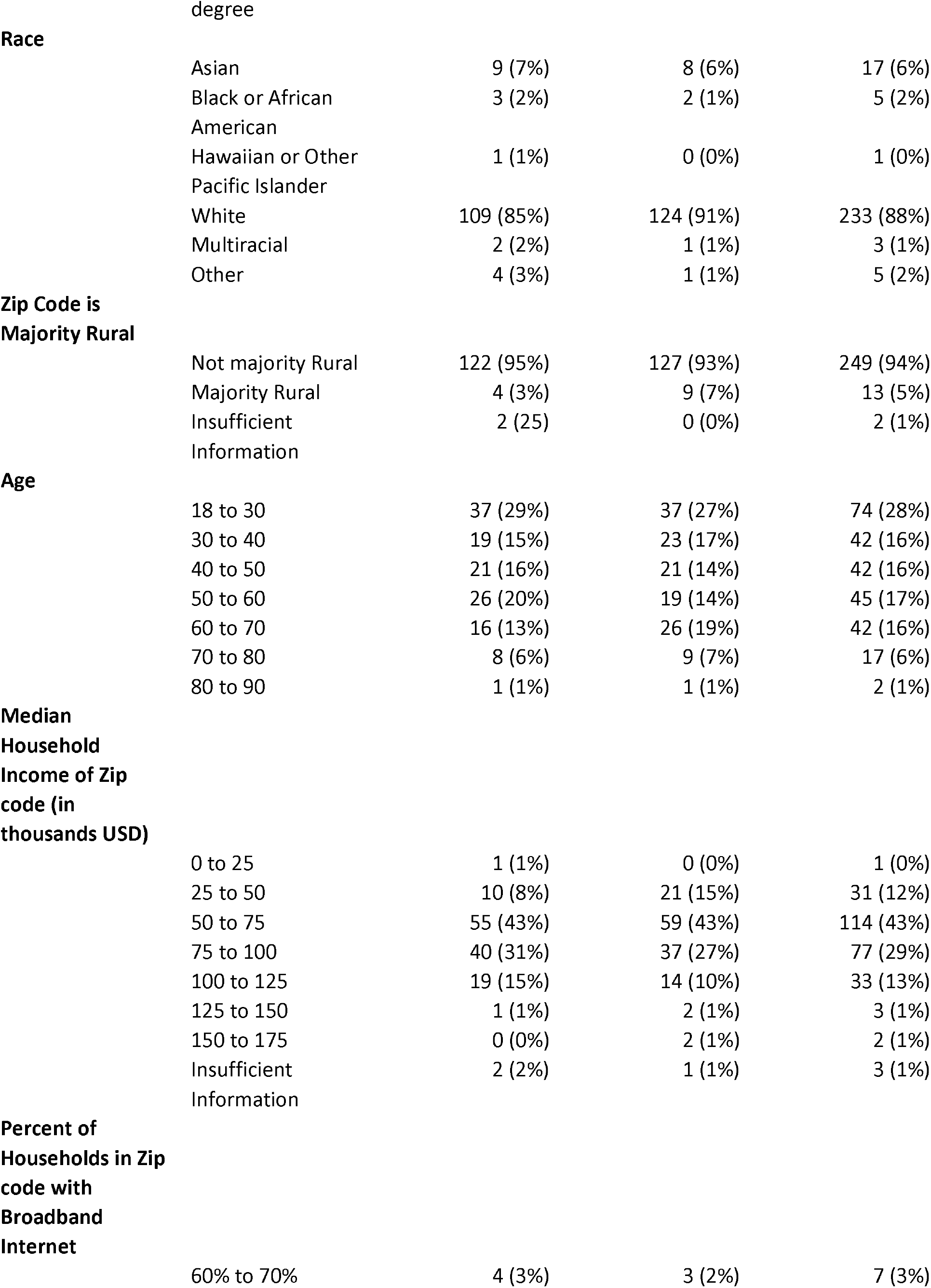
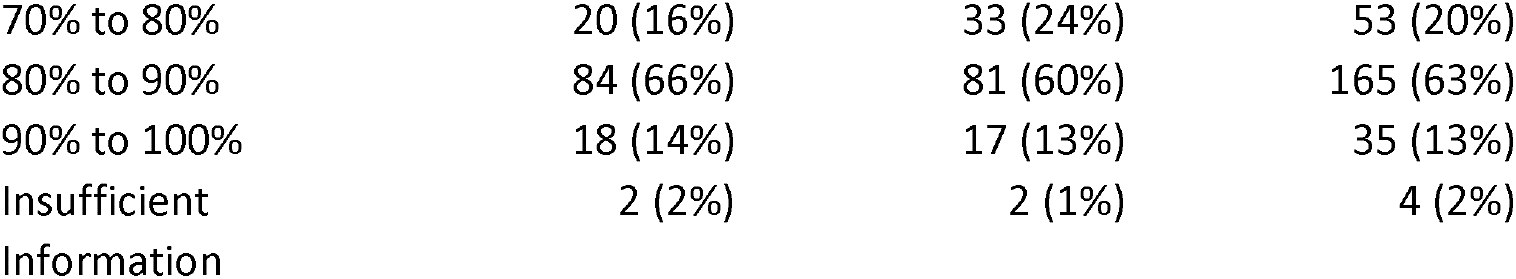
Demographics

|  |  | Survey 1 | Survey 2 | Total |
| --- | --- | --- | --- | --- |
|  |  | 128 | 136 | 264 |
| Gender | Female | 70 (55%) | 84 (62%) | 154 (58%) |
|  | Male | 58 (45%) | 52 (38%) | 110 (42%) |
| Education | High school diploma or GED | 6 (5%) | 7 (5%) | 13 (5%) |
|  | Associate's degree | 9 (7%) | 9 (7%) | 18 (7%) |
|  | Some college | 22 (17%) | 25 (18%) | 47 (18%) |
|  | Bachelor's degree | 50 (39%) | 53 (39%) | 103 (39%) |
|  | Graduate or professional degree | 41(32%) | 42 (31%) | 83 (31%) |
| Race | Asian | 9 (7%) | 8 (6%) | 17 (6%) |
|  | Black or African American | 3 (2%) | 2 (1%) | 5 (2%) |
|  | Hawaiian or Other Pacific Islander | 1 (1%) | 0 (0%) | 1 (0%) |
|  | White | 109 (85%) | 124 (91%) | 233 (88%) |
|  | Multiracial | 2 (2%) | 1 (1%) | 3 (1%) |
|  | Other | 4 (3%) | 1 (1%) | 5 (2%) |
| Zip Code is Majority Rural | Not majority Rural | 122 (95%) | 127 (93%) | 249 (94%) |
|  | Majority Rural | 4 (3%) | 9 (7%) | 13 (5%) |
|  | Insufficient Information | 2 (2%) | 0 (0%) | 2 (1%) |
| Age | 18 to 30 | 37 (29%) | 37 (27%) | 74 (28%) |
|  | 30 to 40 | 19 (15%) | 23 (17%) | 42 (16%) |
|  | 40 to 50 | 21 (16%) | 21 (14%) | 42 (16%) |
|  | 50 to 60 | 26 (20%) | 19 (14%) | 45 (17%) |
|  | 60 to 70 | 16 (13%) | 26 (19%) | 42 (16%) |
|  | 70 to 80 | 8 (6%) | 9 (7%) | 17 (6%) |
|  | 80 to 90 | 1 (1%) | 1 (1%) | 2 (1%) |
| Median Household Income of Zip code (in thousands USD) | 0 to 25 | 1 (1%) | 0 (0%) | 1 (0%) |
|  | 25 to 50 | 10 (8%) | 21 (15%) | 31 (12%) |
|  | 50 to 75 | 55 (43%) | 59 (43%) | 114 (43%) |
|  | 75 to 100 | 40 (31%) | 37 (27%) | 77 (29%) |
|  | 100 to 125 | 19 (15%) | 14 (10%) | 33 (13%) |
|  | 125 to 150 | 1 (1%) | 2 (1%) | 3 (1%) |
|  | 150 to 175 | 0 (0%) | 2 (1%) | 2 (1%) |
|  | Insufficient Information | 2 (2%) | 1 (1%) | 3 (1%) |
| Percent of Households in Zip code with Broadband Internet | 60% to 70% | 4 (3%) | 3 (2%) | 7 (3%) |

|  |  |  |  |
| --- | --- | --- | --- |
| 70% to 80% | 20 (16%) | 33 (24%) | 53 (20%) |
| 80% to 90% | 84 (66%) | 81 (60%) | 165 (63%) |
| 90% to 100% | 18 (14%) | 17 (13%) | 35 (13%) |
| Insufficient Information | 2 (2%) | 2 (1%) | 4 (2%) |

**Table 2:**
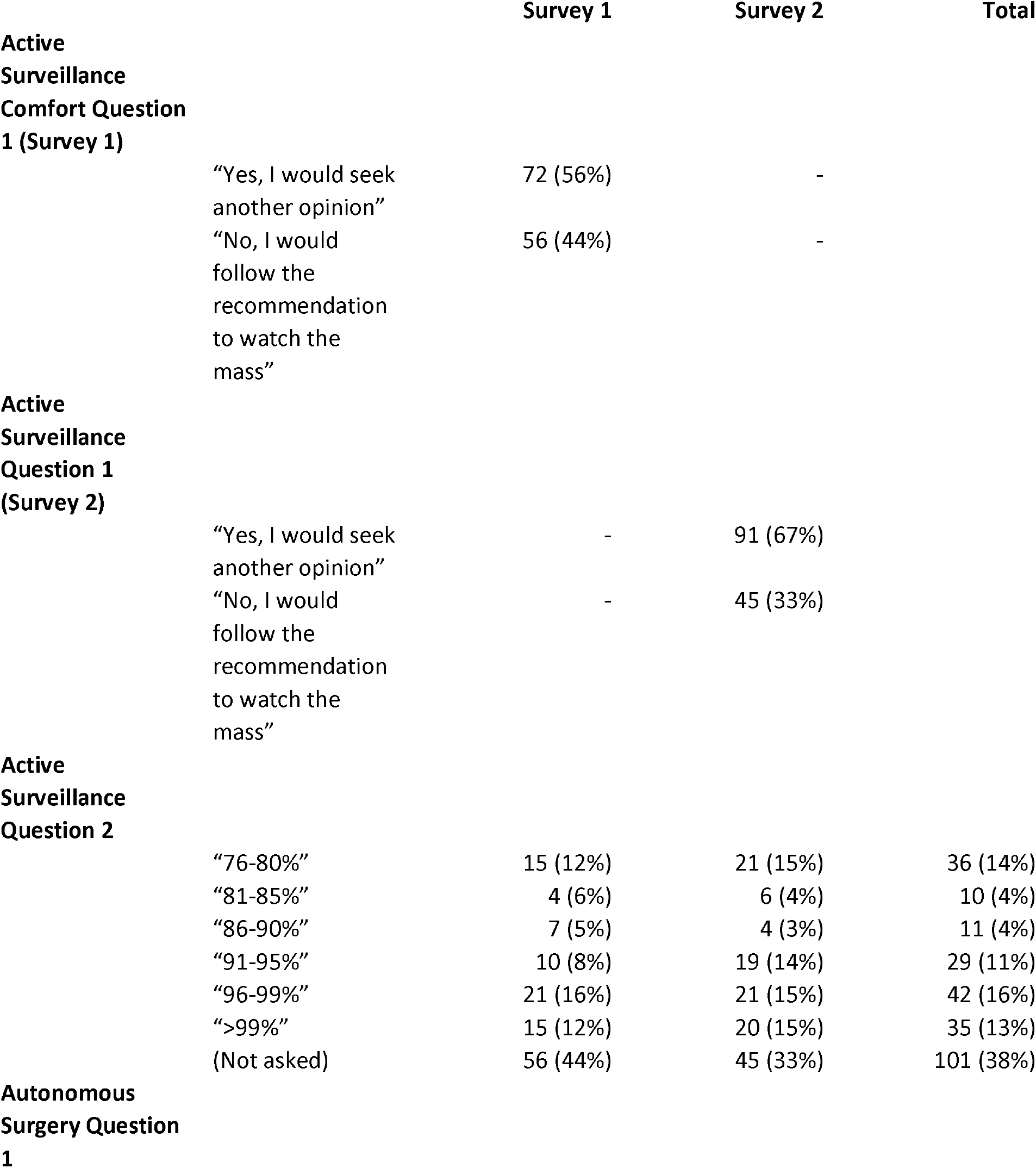
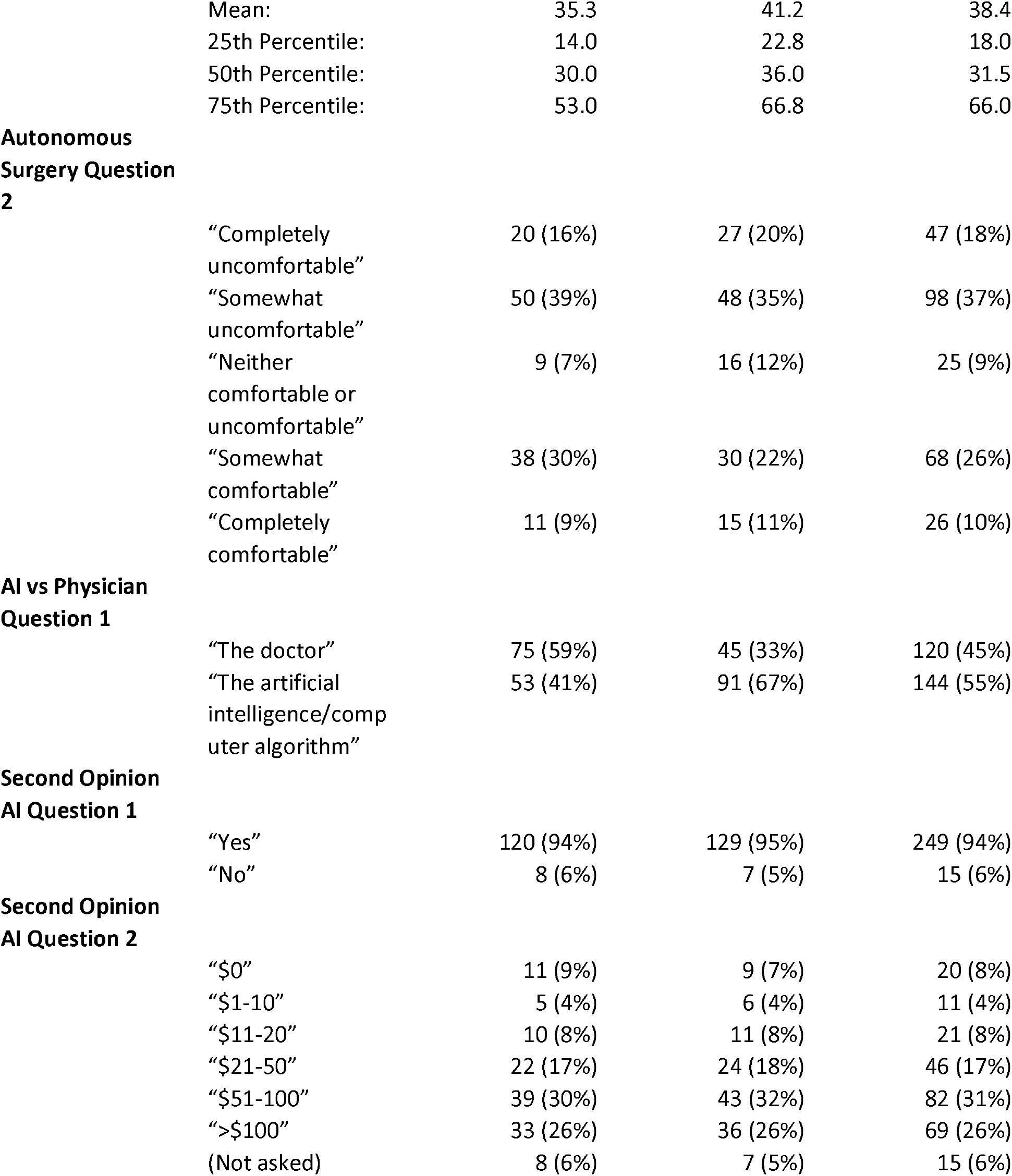
Survey Responses (See Appendix B for complete question text)

|  | Survey 1 | Survey 2 | Total |
| --- | --- | --- | --- |
| **Active Surveillance Comfort Question 1 (Survey 1)** |  |  |  |
| “Yes, I would seek another opinion” | 72 (56%) | - |  |
| “No, I would follow the recommendation to watch the mass” | 56 (44%) | - |  |
| **Active Surveillance Question 1 (Survey 2)** |  |  |  |
| “Yes, I would seek another opinion” | - | 91 (67%) |  |
| “No, I would follow the recommendation to watch the mass” | - | 45 (33%) |  |
| **Active Surveillance Question 2** |  |  |  |
| “76-80%” | 15 (12%) | 21 (15%) | 36 (14%) |
| “81-85%” | 4 (6%) | 6 (4%) | 10 (4%) |
| “86-90%” | 7 (5%) | 4 (3%) | 11 (4%) |
| “91-95%” | 10 (8%) | 19 (14%) | 29 (11%) |
| “96-99%” | 21 (16%) | 21 (15%) | 42 (16%) |
| “>99%” | 15 (12%) | 20 (15%) | 35 (13%) |
| (Not asked) | 56 (44%) | 45 (33%) | 101 (38%) |
| **Autonomous Surgery Question 1** |  |  |  |

|  |  |  |  |  |
| --- | --- | --- | --- | --- |
|  | Mean: | 35.3 | 41.2 | 38.4 |
|  | 25th Percentile: | 14.0 | 22.8 | 18.0 |
|  | 50th Percentile: | 30.0 | 36.0 | 31.5 |
|  | 75th Percentile: | 53.0 | 66.8 | 66.0 |
| **Autonomous Surgery Question 2** |  |  |  |  |
|  | "Completely uncomfortable" | 20 (16%) | 27 (20%) | 47 (18%) |
|  | "Somewhat uncomfortable" | 50 (39%) | 48 (35%) | 98 (37%) |
|  | "Neither comfortable or uncomfortable" | 9 (7%) | 16 (12%) | 25 (9%) |
|  | "Somewhat comfortable" | 38 (30%) | 30 (22%) | 68 (26%) |
|  | "Completely comfortable" | 11 (9%) | 15 (11%) | 26 (10%) |
| **AI vs Physician Question 1** |  |  |  |  |
|  | "The doctor" | 75 (59%) | 45 (33%) | 120 (45%) |
|  | "The artificial intelligence/computer algorithm" | 53 (41%) | 91 (67%) | 144 (55%) |
| **Second Opinion AI Question 1** |  |  |  |  |
|  | "Yes" | 120 (94%) | 129 (95%) | 249 (94%) |
|  | "No" | 8 (6%) | 7 (5%) | 15 (6%) |
| **Second Opinion AI Question 2** |  |  |  |  |
| | "\$0" | 11 (9%) | 9 (7%) | 20 (8%) |
| | "\$1-10" | 5 (4%) | 6 (4%) | 11 (4%) |
| | "\$11-20" | 10 (8%) | 11 (8%) | 21 (8%) |
| | "\$21-50" | 22 (17%) | 24 (18%) | 46 (17%) |
| | "\$51-100" | 39 (30%) | 43 (32%) | 82 (31%) |
| | ">\$100" | 33 (26%) | 36 (26%) | 69 (26%) |
|  | (Not asked) | 8 (6%) | 7 (5%) | 15 (6%) |

### Active Surveillance

Of the 264 responses, 163 individuals (62%) indicated that they would seek a second opinion given the recommendation to wait on a surgery and watch the growth of the mass via imaging and the rest indicated that they would follow the recommendation. Of those that received the version of the question indicating that the recommendation was based on the evaluation of an AI algorithm 67% indicated that they would seek a second opinion compared to 43 % when the recommendation was based on a physician recommendation, however this was not found to be significant in a chi-square test (p = 0.098). In multivariate analysis no demographic features were found to be significant in predicting whether the participant would seek a second opinion.

Of those who indicated that they would seek a second opinion, a plurality of 42 out of 101 (42%), indicated that they would prefer a confidence level between 96-99%. See Table 2 for a complete summary of responses. Lower levels of education were correlated with lower levels of confidence required: (in order increasing absolute value of coefficients) those with some college education (p = 0.032), those with an associate’s degree (p = 0.024) and those with a high school or GED (p = 0.0074) (see Table 3).

**Table 3:**
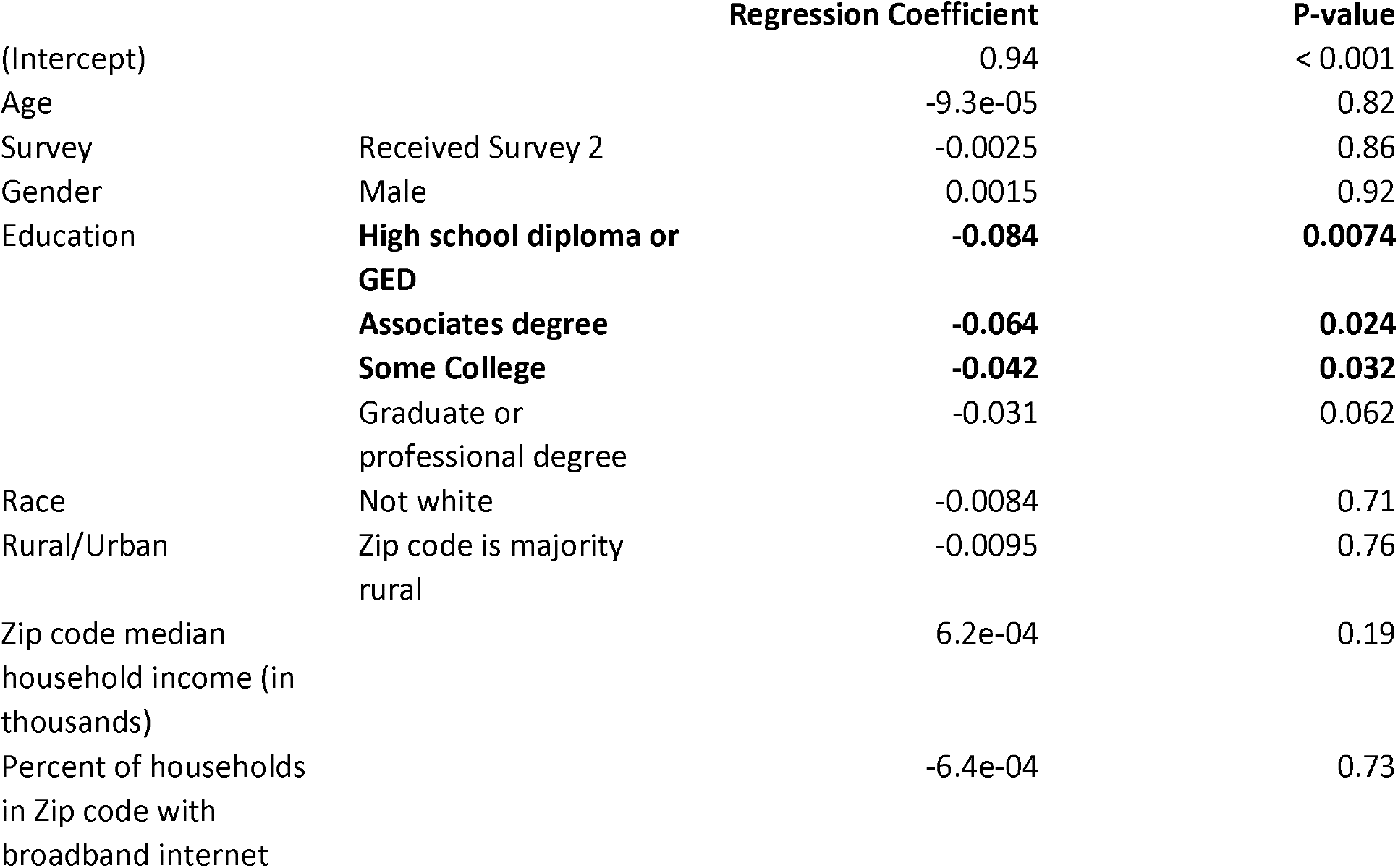
Multivariate regression of percentage confidence required in order to feel comfortable with active surveillance

|  |  | Regression Coefficient | P-value |
| --- | --- | --- | --- |
| (Intercept) |  | 0.94 | < 0.001 |
| Age |  | -9.3e-05 | 0.82 |
| Survey | Received Survey 2 | -0.0025 | 0.86 |
| Gender | Male | 0.0015 | 0.92 |
| Education | **High school diploma or GED** | **-0.084** | **0.0074** |
|  | **Associates degree** | **-0.064** | **0.024** |
|  | **Some College** | **-0.042** | **0.032** |
|  | Graduate or professional degree | -0.031 | 0.062 |
| Race | Not white | -0.0084 | 0.71 |
| Rural/Urban | Zip code is majority rural | -0.0095 | 0.76 |
| Zip code median household income (in thousands) |  | 6.2e-04 | 0.19 |
| Percent of households in Zip code with broadband internet |  | -6.4e-04 | 0.73 |

### Autonomous Surgery

Despite the fact that there are no FDA approved autonomous robotic surgical systems in the United States, when asked to estimate the percentage of a standard robotic surgery that was autonomous, participants selected values ranging from 0% to 100%. The median value selected was 31.5% and the mean 38.4%. No demographics were found to be significant in multiple regression against this estimation.

With regard to comfort with robotic surgery, 47 participants (18%) indicated that they were ‘completely uncomfortable’ with robotic surgery, 98 (37%) ‘somewhat uncomfortable’, 25 (9%) ‘neither comfortable or uncomfortable’, 68 (26%) ‘somewhat comfortable’ and 26 (10%) ‘completely comfortable’. Results of multiple regression can be found in Table 4. Older respondents and those whose zip codes had a higher percentage of households with a broadband internet subscription were significantly more likely to choose ‘completely uncomfortable’ vs other responses. The odds ratio of age was 1.03 (p = 0.0061) and percentage of households with broadband internet had odds ratio 1.13 (p = 0.032). Those who indicated zip codes with higher median incomes were much less likely to select ‘completely uncomfortable’, with odds ratio 0.57 (p = 0.00078). Higher median incomes were more likely to select ‘Somewhat comfortable’ vs other options (OR = 1.03, p = 0.014). Being male (OR = 5.02, p = 0.0010) and having an associate’s degree (OR = 4.36, p = 0.037) were significant among those selecting ‘Completely comfortable’ vs all other options.

**Table 4:**
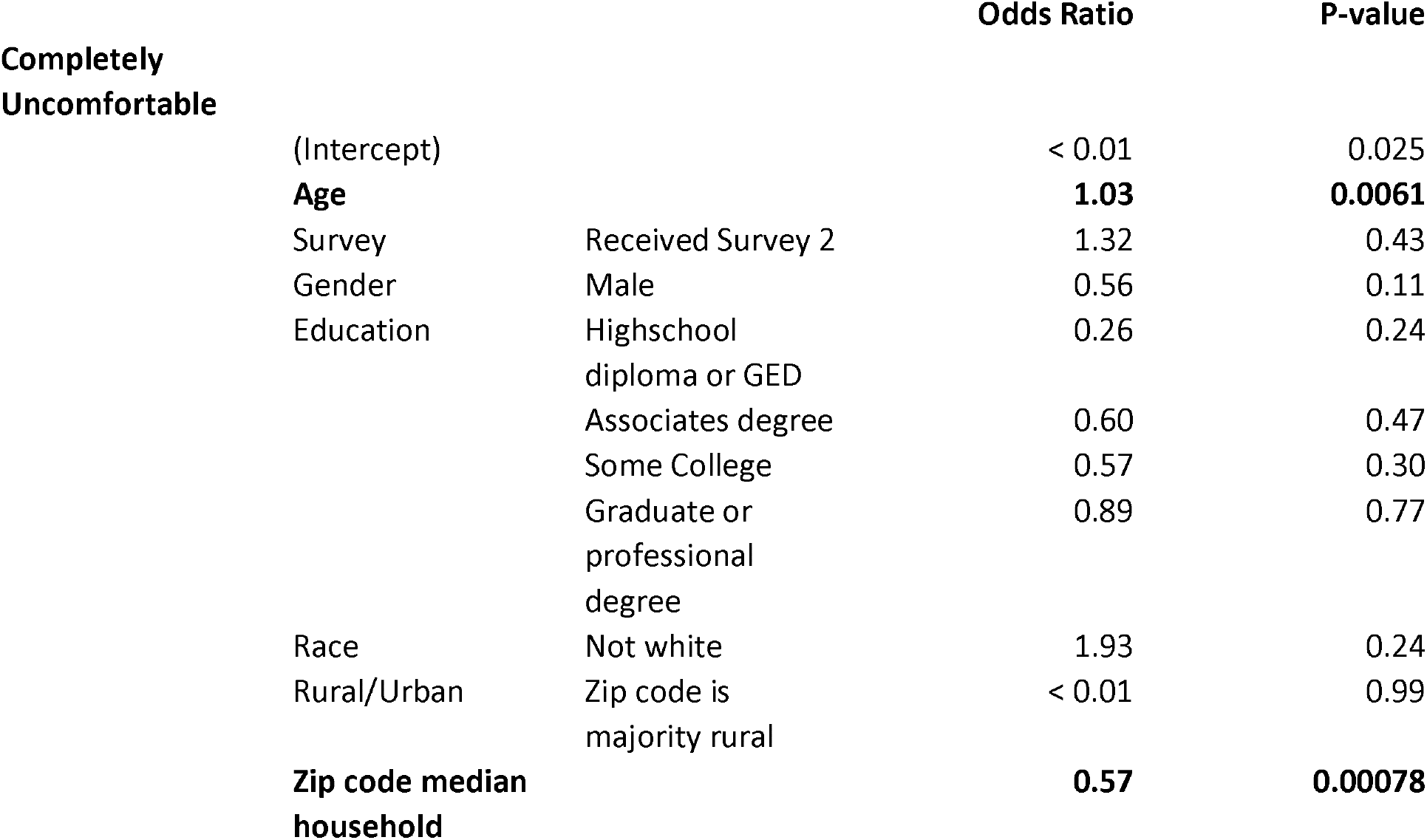
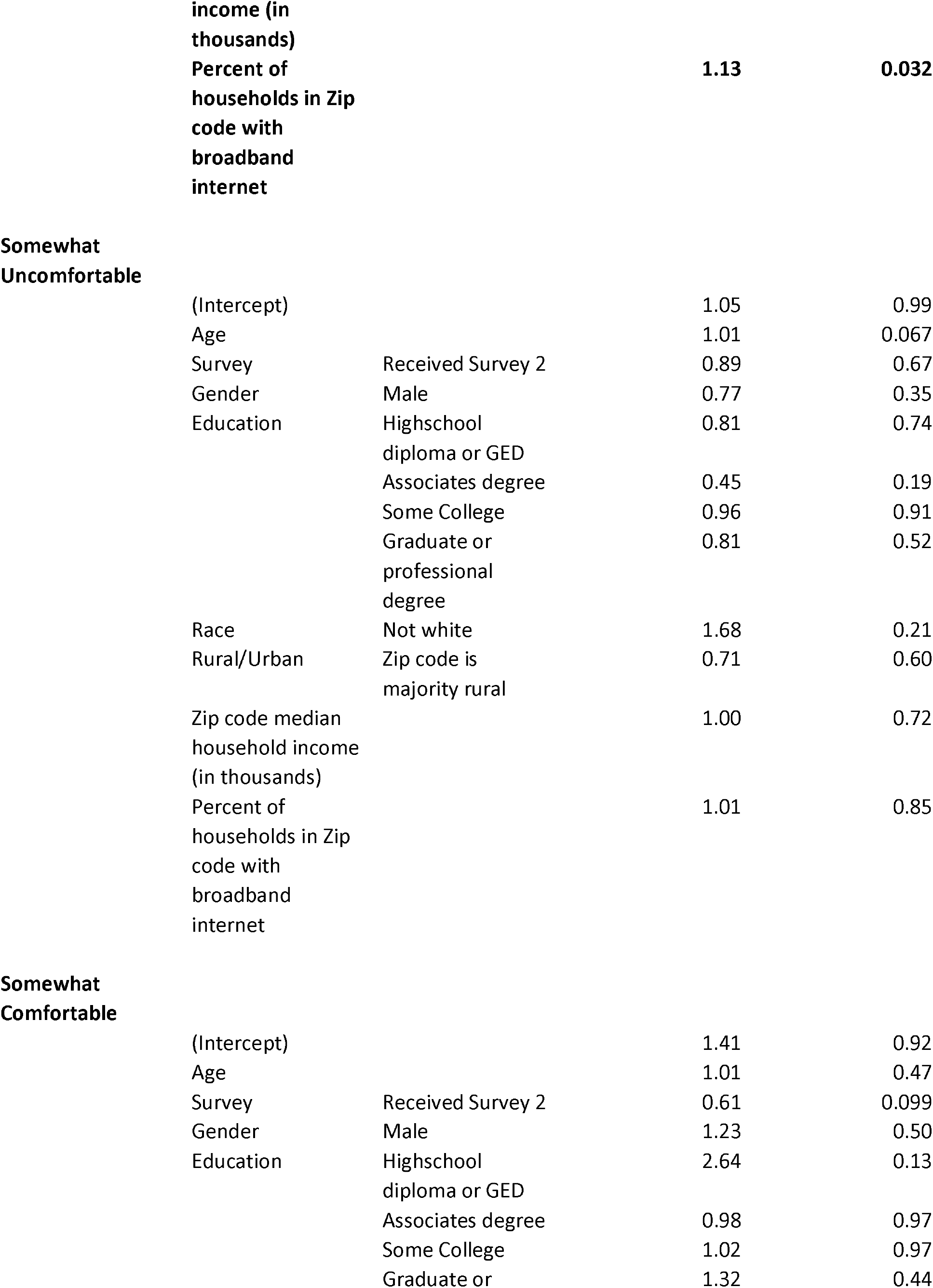
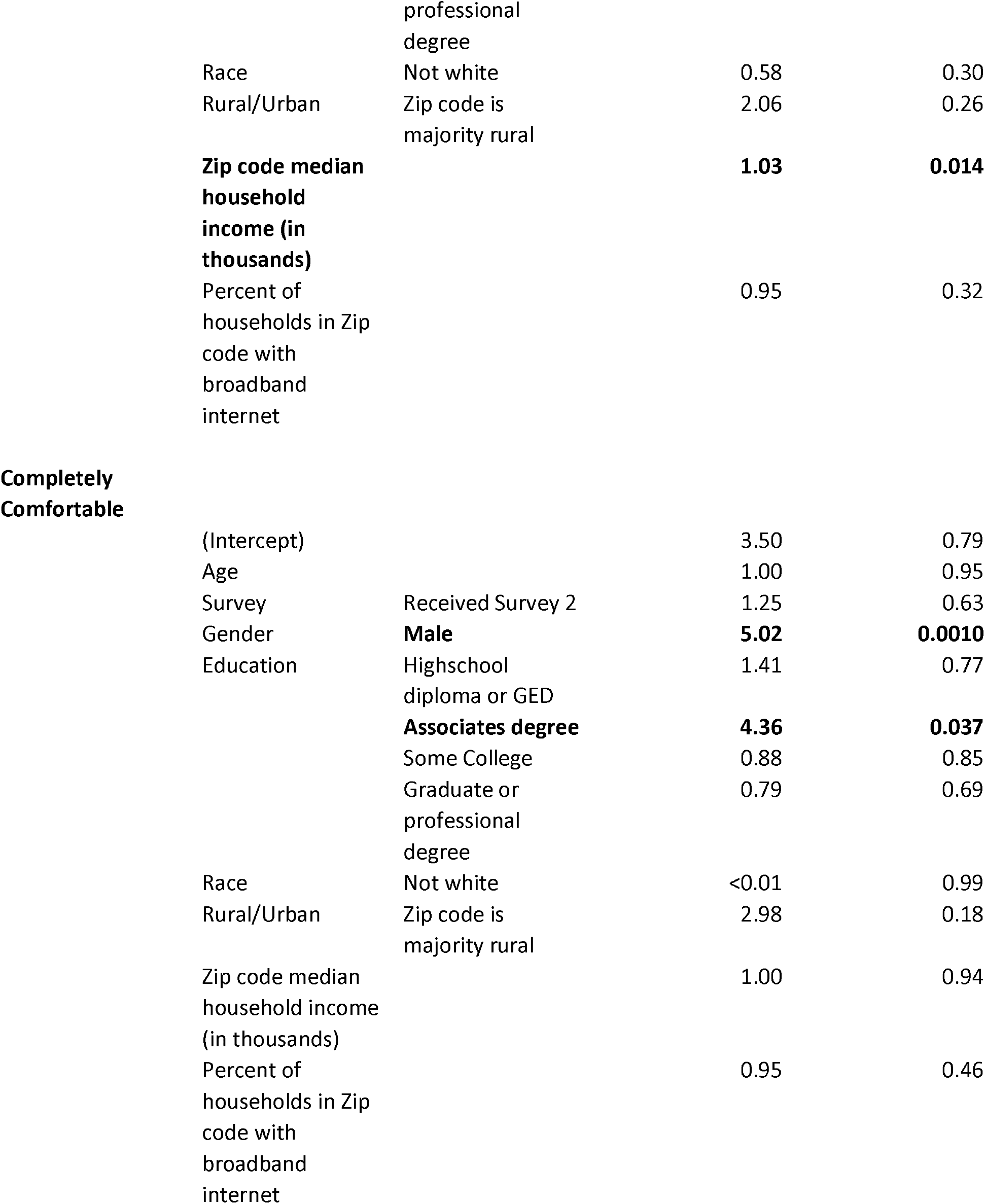
Multivariate logistic regression of each level of comfort with robotic surgery

Taken together as a continuous measure of comfort (ranging from 1-5) being male (p = 0.0017), having an associate’s degree (p = 0.041) and indicating a zip code with a higher median income (p = 0.0012) all again had a positive relationship with comfort with robotic surgery. Selecting a race other than white (p= 0.014) and neighborhoods with higher number of households with a broadband internet subscription (p= 0.035) were both negatively related to comfort with robotic surgery.

### AI vs Physician

Given the scenario that a physician and an AI algorithm disagree in their estimation that a mass is benign (the doctor believing it to be cancerous and the AI believing it to be benign) roughly half of respondents, 144 out of 264, indicate a higher confidence in the AI. Those who randomly received the second version of the survey, where an earlier question suggested AI can provide recommendations, were much more likely to trust the AI’s diagnosis over the doctor (OR = 3.45, p = <0.001). The majority of participants who received the first survey had greater confidence in the doctor’s diagnosis while the reverse was true for those who received the second, as highlighted in Figure 1. No other predictors were found to be significant. Complete multiple regression results can be found in Table 5.

**Table 5:** Multivariate logistic regression of preference for a physician recommendation over and AI

|  |  | Odds Ratio | P-value |
| --- | --- | --- | --- |
| (Intercept) |  | 1090 | 0.022 |
| Age |  | 1.00 | 0.81 |
| **Survey** | **Received survey 2** | **0.29** | **< 0.001** |
| Gender | Male | 1.18 | 0.55 |
| Education | High school diploma or GED | 2.38 | 0.20 |
|  | Associates degree | 0.61 | 0.37 |
|  | Some college | 1.22 | 0.60 |
|  | Graduate or professional degree | 0.64 | 0.18 |
| Race: | Not white | 0.81 | 0.63 |
| Urban/Rural | Zip code is majority rural | 2.20 | 0.24 |
| Zip code median household income (in thousands) |  | 1.00 | 0.69 |
| Percent of households in Zip code with broadband internet |  | 0.92 | 0.054 |

**Figure 1:**
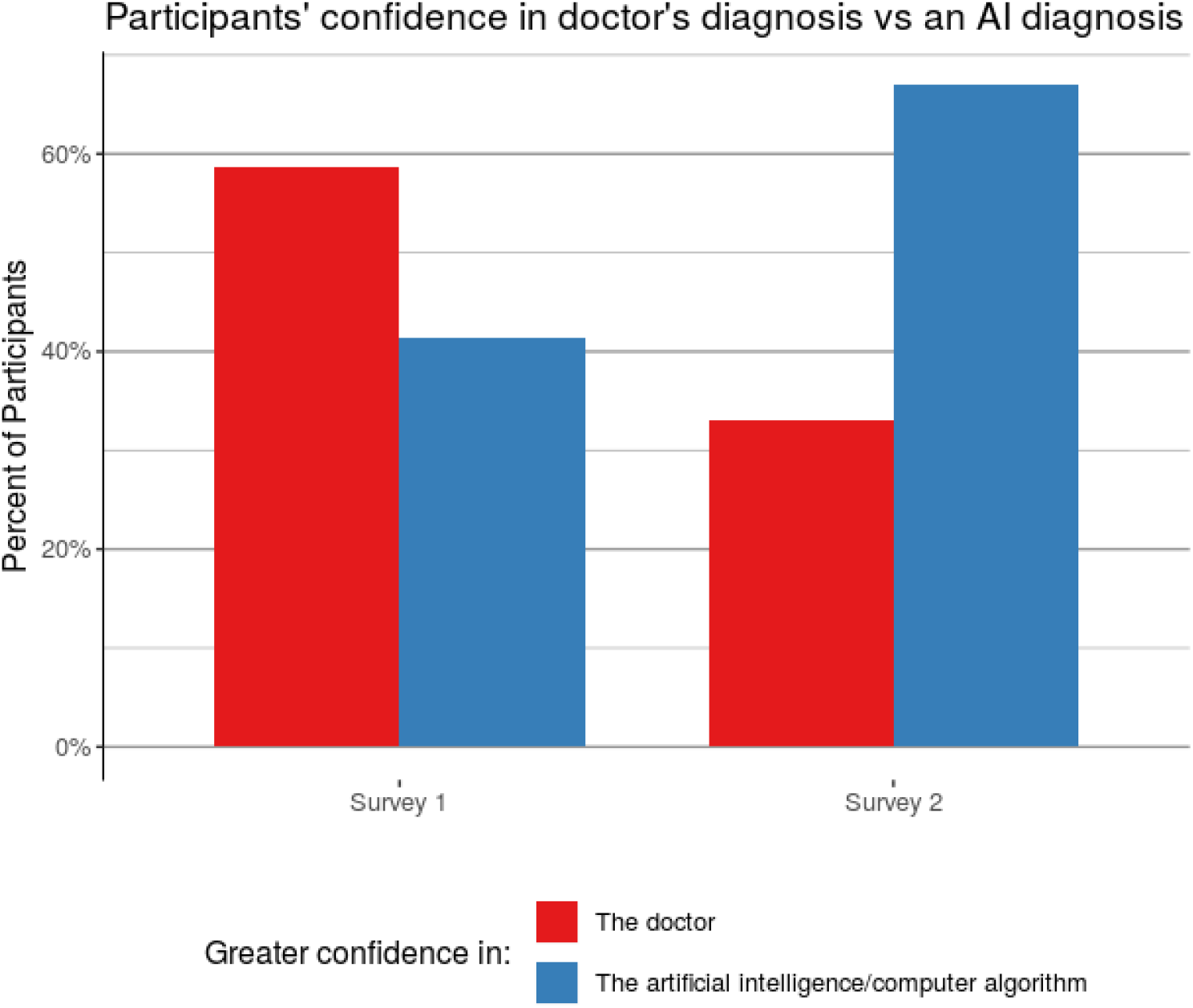
AI vs Physician Confidence. Percent of participants in each survey that selected greater confidence in either the doctor’s diagnosis or an AI diagnosis given the doctor (See Appendix B for full question text)

### Second Opinion AI

Almost all participants (94%) indicated that they would be comfortable with sending their medical imaging to an AI algorithm for review via a website. Of those individuals, 92% indicated that they would be willing to pay for such a service.From lowest to highest: 11 (4%) selected ‘$1-10’, 21 (8%) selected ‘$11-20’, 46 (18%) selected ‘$21-50’, 82 (33%) selected ‘$51-100’ and 69 (28%) indicated they would be willing to pay more than $100. No significant relationships were found between demographic variables and individuals’ responses in this category.

## Discussion

The results of this survey show a great diversity in opinions on AI and robotic surgery in the diagnosis and treatment of cancer. We did not find evidence to support a relationship with many demographic features such as age, gender, rural vs. urban for most questions. Lower levels of education were correlated with lower certainty required in the benign nature of a mass in order to be comfortable with waiting on surgery. A number of factors were significantly associated with comfort with robotic surgery, including lower age, Caucasian race, lack of broadband internet connections, higher median income areas, having only an associate’s degree, and male gender. The most notable relationship found in this survey was the positive association between receiving the version of the survey that posited an AI estimate in the first question and having higher confidence in the diagnosis of an AI when it disagreed with the physician. These results suggest an optimism toward medical applications of AI, but poor understanding of its current capabilities and use.

In spite of the extensive adoption of robotic surgery in the US, there is still significant misunderstanding about its nature. Even though surgical robots do not currently operate autonomously in standard practice, 88% of participants believe that it does, at least to some degree. While our sample is a convenience sample, individuals of every race age and education overestimated the presence of autonomous behavior.

Also, in contrast with initial customer surveys [6] we found evidence to suggest a certain openness to artificial intelligence applications. This finding aligns with recent research suggesting that public perception and optimism of AI as a whole has risen markedly [11]. A third of all participants believed that robotic surgery contained some autonomous behavior and indicated that they were either ‘Somewhat comfortable’ or ‘Completely comfortable’ with such a procedure.

Almost a third of individuals who received a recommendation to wait and monitor a tumor’s growth based on an AI estimation were comfortable with following the recommendation. Many patients could feel that an AI gives them additional certainty in their diagnosis which has been shown to be predictive of quality of life in individuals undergoing monitoring of small renal tumors [12].

The significant priming effect of first question suggests a sensitivity in overall opinion to how an AI diagnosis is presented. Those that receive an indication that an AI can make a diagnosis show greater confidence in it. Given the widespread research into AI applications in medicine, there has been significant media coverage. In May 2019, the New York Times published an article headlined “A.I. Took a Test to Detect Lung Cancer. It Got an A” [13]. Recently Google AI announced a partnership with the Mayo Clinic [14]. Such media coverage has the possibility of increasing the public’s confidence in and perhaps inflates the capabilities of such technology.

Given public optimism about AI and the significant confusion about current technological capabilities, there is a great need for careful patient counseling. There is unquestionably a large number of promising applications for artificial intelligence in medicine, but current implementations of AI are scarce and media representations of AI are often far beyond what is currently robust in practice, which likely contributes to the optimism that we observe. This optimism might be beneficial for early adoption, but the associated misperceptions could hinder informed decision making and eventually damage trust. It’s therefore important to discuss with patients the role that technology is playing in their care, along with its limitations.

## Conclusion

There is significant variance in population perception and understanding of AI and robotic surgery. Proactive conversations with patients who might be undergoing robotic surgery may be beneficial. Ongoing research at both commercial and academic institutions, technology companies and medical institutions, inevitably spurs media attention on medical AI. Our results suggest that individuals’ opinions and confidence in AI are sensitive to such representations, and if patients overestimate the capabilities of AI in medicine, it could lead them to take bigger risks and accept greater uncertainties than they might be otherwise comfortable with. In addition, an initial overestimation of AI capabilities may lead to early disappointment causing techniques with real potential to be abandoned due to unmet expectations. As research continues in AI applications, there is a need for clear communication going forward about the strengths and limitations of such techniques.

### Sources of Support

Research reported in this publication was supported by the National Cancer Institute of the National Institutes of Health under Award Number R01CA225435. The content is solely the responsibility of the authors and does not necessarily represent the official views of the National Institutes of Health.

Additional support was provided by the Climb for Kidney Cancer Foundation.

## Data Availability

The full survey responses are available to researchers upon reasonable request.

## Appendix A: Tables and Figures

## Appendix B: Survey Text

Questions and responses for Survey 1 are as follows.

### Questions on Active Surveillance

“Suppose you begin to experience stomach pain and decide to visit a medical clinic. Your doctor orders an abdominal CT scan which shows a 3 cm (1.5 inch) mass that could be a kidney cancer (see image below). She then refers you to a urologist for a consult for surgical removal of the mass, which would have a 99% chance of cure if it is cancer. The urologist explains that masses with CT characteristics like yours are unlikely to be cancerous, estimating the chance this mass is a kidney cancer is only 25%. He suggests that you undergo follow-up imaging every six months moving forward, and that if the mass changes considerably he will surgically remove it. Would you seek a second opinion?”

“Yes, I would seek another opinion”,

“No, I would follow the recommendation to watch the mass”

“If you feel uncomfortable not treating a mass with a 25% chance of cancer, what level of certainty do you need to have before your feel comfortable not treating a mass?”

“76-80%”, “81-85%”, “86-90%”, “91-95%”, “96-99%”, “>99”

### Autonomous Surgery

“Today, many surgical procedures are done with the assistance of a robot. In the average robotic assisted surgery, what proportion of the procedure do you estimate that the robot is operating autonomously (or in other words, by itself without guidance from the surgeon)? “

(User input slider 0-100)

“If a robot were developed to perform a cancer surgery completely autonomously (in other words, the robot performs the surgery by itself without guidance from a surgeon). What would your comfort level be to undergo that type of operation?”

“Completely uncomfortable”, “Somewhat uncomfortable”, “Neither comfortable or uncomfortable”, “Somewhat comfortable”, “Completely comfortable”

### AI vs Physician

“Suppose you visit your clinic for nausea and your doctor orders a CT scan of the abdomen. Your doctor then reviews your imaging and doesn’t find an explanation for your nausea, but unexpectedly finds an adrenal mass which she believes is cancer. However, an artificial intelligence computer algorithm also reviews your imaging and concludes your adrenal mass is benign. In which diagnosis do you have a higher confidence? “

“The doctor”, “The artificial intelligence/computer algorithm”

### Second Opinion AI

“If there was a website where an artificial intelligence/computer algorithm was able to read your CT scan to give you a second opinion on whether a mass is cancer or not, would you allow your CT scan to be read by a computer?”

“Yes”, “No”

“If you would allow a computer to read your CT scan how much would you be willing to pay for that service?”

“$0”, “$1-10”, “$11-20”, “$21-50”, “$51-100”, “>$100”

### Survey 2

#### Questions on Active Surveillance

“Suppose you begin to experience stomach pain and decide to visit a medical clinic. Your doctor orders an abdominal CT scan which shows a 3 cm (1.5 inch) mass that could be a kidney cancer (see image below). She then refers you to a urologist for a consult for surgical removal of the mass, which would have a 99% chance of cure if it is cancer. The urologist explains that an artificial intelligence/computer algorithm concluded that masses with CT characteristics like yours are unlikely to be cancerous, estimating the chance this mass is a kidney cancer is only 25%. He suggests that you undergo follow-up imaging every six months moving forward, and that if the mass changes considerably he will surgically remove it. Would you seek a second opinion?”

“Yes, I would seek another opinion”,

“No, I would follow the recommendation to watch the mass”

“If you feel uncomfortable not treating a mass with a 25% chance of cancer, what level of certainty do you need to have before your feel comfortable not treating a mass?”

“76-80%”, “81-85%”, “86-90%”, “91-95%”, “96-99%”, “>99”

#### Autonomous Surgery

“Today, many surgical procedures are done with the assistance of a robot. In the average robotic assisted surgery, what proportion of the procedure do you estimate that the robot is operating autonomously (or in other words, by itself without guidance from the surgeon)? “

(User input slider 0-100)

“If a robot were developed to perform a cancer surgery completely autonomously (in other words, the robot performs the surgery by itself without guidance from a surgeon). What would your comfort level be to undergo that type of operation?”

“Completely uncomfortable”, “Somewhat uncomfortable”, “Neither comfortable or uncomfortable”, “Somewhat comfortable”, “Completely comfortable”

#### AI vs Physician

“Suppose you visit your clinic for nausea and your doctor orders a CT scan of the abdomen. Your doctor then reviews your imaging and doesn’t find an explanation for your nausea, but unexpectedly finds an adrenal mass which she believes is cancer. However, an artificial intelligence computer algorithm also reviews your imaging and concludes your adrenal mass is benign. In which diagnosis do you have a higher confidence? “

“The doctor”, “The artificial intelligence/computer algorithm”

#### Second Opinion AI

“If there was a website where an artificial intelligence/computer algorithm was able to read your CT scan to give you a second opinion on whether a mass is cancer or not, would you allow your CT scan to be read by a computer?”

“Yes”, “No”

“If you would allow a computer to read your CT scan how much would you be willing to pay for that service?”

“$0”, “$1-10”, “$11-20”, “$21-50”, “$51-100”, “>$100”

## References

[1] James H. Thrall, Xiang Li, Quanzheng Li, Cinthia Cruz, Synho Do, Keith Dreyer, and James Brink. Artificial intelligence and machine learning in radiology: Opportunities, challenges, pitfalls, and criteria for success. 15(3):504–508. ISSN 15461440. doi: 10.1016/j.jacr.2017.12.026. URL https://linkinghub.elsevier.com/retrieve/pii/S154614401731671X.

[2] Chayakrit Krittanawong, HongJu Zhang, Zhen Wang, Mehmet Aydar, and Takeshi Kitai. Artificial intelligence in precision cardiovascular medicine. 69(21):2657–2664. ISSN 0735-1097. doi: 10.1016/j.jacc.2017.03.571. URL http://www.sciencedirect.com/science/article/pii/S0735109717368456.

[3] Andrea Cestari. Predictive models in urology. 80(1):42–45. ISSN 0391-5603. doi: 10.5301/RU.2013.10744. URL https://doi.org/10.5301/RU.2013.10744.

[4] Jong Kyu Choi and Yong Gu Ji. Investigating the importance of trust on adopting an autonomous vehicle. 31(10):692–702. ISSN 1044-7318. doi: 10.1080/10447318.2015.1070549. URL https://doi.org/10.1080/10447318.2015.1070549.

[5] Clemence Cavoli, Brian Phillips, Tom Cohen, and Peter Jones. Social and behavioural questions associated with automated vehicles: A literature review. page 124.

[6] Chiara Longoni, Andrea Bonezzi, and Carey K. Morewedge. Resistance to medical artificial intelligence. doi: 10.1093/jcr/ucz013. URL https://academic.oup.com/jcr/advance-article/doi/10.1093/jcr/ucz013/5485292.

[7] Jens J. Rassweiler, Riccardo Autorino, Jan Klein, Alex Mottrie, Ali Serdar Goezen, Jens-Uwe Stolzenburg, Koon H. Rha, Marc Schurr, Jihad Kaouk, Vipul Patel, Prokar Dasgupta, and Evangelos Liatsikos. Future of robotic surgery in urology. 120(6):822–841. ISSN 1464-410X. doi: 10.1111/bju.13851.

[8] Arif Ahmad, Zoha F. Ahmad, Jared D. Carleton, and Ashish Agarwala. Robotic surgery: current perceptions and the clinical evidence. 31(1):255–263. ISSN 1432-2218. doi: 10.1007/s00464-016-4966-y. URL https://doi.org/10.1007/s00464-016-4966-y.

[9] Joshua A. Boys, Evan T. Alicuben, Michael J. DeMeester, Stephanie G. Worrell, Daniel S. Oh, Jeffrey A. Hagen, and Steven R. DeMeester. Public perceptions on robotic surgery, hospitals with robots, and surgeons that use them. 30(4):1310–1316. ISSN 1432-2218. doi: 10.1007/s00464-015-4368-6. URL https://doi.org/10.1007/s00464-015-4368-6.

[10] American Community Survey Office. 2017 data profiles. URL https://www.census.gov/acs/www/data/data-tables-and-tools/data-profiles/.

[11] Ethan Fast and Eric Horvitz. Long-term trends in the public perception of artificial intelligence. URL http://arxiv.org/abs/1609.04904.

[12] Patricia A. Parker, Frances Alba, Bryan Fellman, Diana L. Urbauer, Yisheng Li, Jose A. Karam, Nizar Tannir, Eric Jonasch, Christopher G. Wood, and Surena F. Matin. Illness uncertainty and quality oflife of patients with small renal tumors undergoing watchful waiting: A 2-year prospective study. 63 (6):1122–1127. ISSN 0302-2838. doi: 10.1016/j.eururo.2013.01.034. URL http://www.sciencedirect.com/science/article/pii/S0302283813001036.

[13] Denise Grady. A.i. took a test to detect lung cancer. it got an a. ISSN 0362-4331. URL https://www.nytimes.com/2019/05/20/health/cancer-artificial-intelligence-ct-scans.html.

[14] Mayo clinic selects google as strategic partner for health care innovation, cloud computing. URL https://newsnetwork.mayoclinic.org/discussion/mayo-clinic-selects-google-as-strategic-partner-for-health-care-innovation-cloud-computing/.

